# Long COVID risk and severity after COVID-19 infections and reinfections in Quebec healthcare workers

**DOI:** 10.1101/2025.05.08.25327059

**Authors:** Sara Carazo, Manale Ouakki, Nektaria Nicolakakis, Emilia Liana Falcone, Danuta M Skowronski, Marie-José Durand, Marie-France Coutu, Simon Décary, Isaora Z Dialahy, Olivia Drescher, Elisabeth Canitrot, Carrie Anna McGinn, Philippe Latouche, Robert Laforce, Clemence Dallaire, Geoffroy Denis, Alain Piché, Gaston De Serres

**Affiliations:** Social and preventive medicine department, Faculty of medicine, Laval University, Quebec City, Quebec, Canada; Centre Hospitalier Universitaire (CHU) de Québec–Université Laval Research Center, Quebec City, Quebec, Canada; Department of biological risks. National Institute of Public Health of Quebec. Quebec City, Quebec, Canada; Department of Environmental and Occupational Health and of Toxicology. National Institute of Public Health of Quebec, Montreal, Quebec, Canada; Center for Immunity, Inflammation and Infectious Diseases, Montreal Clinical Research Institute, Montreal, Quebec, Canada; Department of Medicine, Faculty of Medicine, Université de Montréal, Montreal, Quebec, Canada; Immunization Programs and Vaccine Preventable Diseases Services, BC Centre for Disease Control, Vancouver, British Columbia, Canada; School of Rehabilitation, Université de Sherbrooke, Faculty of Medicine and Health Sciences, Sherbrooke, QC, Canada; Hôpital Charles-Le Moyne Research Center, Longueuil, Quebec, Canada; Centre intégré universitaire de santé et de services sociaux de l’Estrie-Centre hospitalier universitaire de Sherbrooke, Sherbrooke, Quebec, Canada; Office of Information and Studies on Population Health, National Institute of Public Health of Quebec, Quebec City, Quebec, Canada; Patient-research partner; Faculty of nursing, Laval University, Quebec City, Quebec, Canada; Ministry of Health and Social Services, Quebec City, Quebec, Canada; School of Population and Global Health, McGill University, Montreal, Quebec, Canada

## Abstract

**Importance:** Long COVID, a chronic condition following SARS-CoV-2 infection, affects millions of people worldwide and can lead to significant functional impairment. Estimates of long COVID risk after a first COVID-19 infection vary, and data on risk following reinfections remain lacking.

**Objective:** To estimate and compare long COVID risk and severity with first COVID-19 infection versus reinfections among healthcare workers (HCWs).

**Design:** Retrospective cohort study based on an electronic survey among Quebec HCWs conducted between May 16 and June 15, 2023. A short telephone survey among randomly selected non-respondents further assessed potential response bias.

**Setting:** Population-based study in Quebec, Canada.

**Participants:** 397 222 HCWs were invited to participate in the electronic survey and 10 500 in the telephone survey.

**Main outcomes and measures:** We defined long COVID cases as HCWs self-reporting COVID-19-attributed symptoms lasting ≥12 weeks, classified as mild, moderate, or severe based on perceived symptom intensity. We compared self-reported symptoms and functional limitations of cases to COVID controls (infected participants without long COVID) and non-COVID controls (uninfected participants). Risk and prevalence were estimated by number of infections, likely infecting variant and perceived acute COVID-19 severity. We conducted symptom clustering analyses using unsupervised learning techniques.

**Results:** Estimated long COVID risk following any COVID-19 infection was similar among 22 496 online survey participants (17.0% [95%CI, 16.3%–17.6%] and 3 978 telephone survey participants (15.9% [14.6%–17.2%]. The cumulative risk increased with the number of infections, but reinfections were associated with three times lower risk of long COVID than first infections.

Pre-Omicron infections and severe acute COVID-19 episodes correlated with higher long COVID risk and severity. Among prevalent long COVID cases, 43% were moderate and 33% severe. Compared to controls, dyspnea, neurocognitive symptoms, post-exertional malaise and smell/taste disturbances were most strongly associated with long COVID. Cluster analysis identified seven symptom groups with systemic, neurocognitive, pulmonary, and mood-related clusters being the most prevalent. Severe long COVID cases exhibited multiple symptom clusters and greater functional limitations.

**Conclusions:** Long COVID is a common and disabling condition among HCWs. Societal and healthcare burden remains important and will continue to accrue given ongoing SARS-CoV-2 transmission and long COVID risk with reinfections.

**KEY POINTS:** *Question:* Does the risk and severity of long COVID differ for first COVID-19 infections versus reinfections among healthcare workers in Quebec, Canada?

*Findings:* The risk of long COVID was ∼15% with first COVID-19 infection, but three-fold lower with reinfections. Risk was higher with COVID-19 infections due to ancestral strains and with more severe acute episodes. Long COVID risk and severity compromised self-rated health, physical capacity and cognitive function.

*Meaning:* Long COVID is a common and disabling condition among workers. Societal and healthcare burden remains important and will continue to accrue given ongoing SARS-CoV-2 transmission and long COVID risk with reinfections.

## INTRODUCTION

Long COVID, also known as post-COVID-19 condition, is an infection-associated chronic multi-systemic condition, with symptoms lasting at least three months, and presenting as a continuous, relapsing and remitting, or progressive disease state.^1^ Affected individuals often experience significant functional impairment, as well as emotional and physical distress. It is estimated that over 400 million people worldwide suffer from long COVID.^2,3^ While long COVID risk after a first COVID-19 episode varies from 10% to 70% for ambulatory and hospitalized acute cases,^4,5^ data on long COVID risk and severity following reinfections are limited.^6^

In the absence of specific diagnostic biomarkers, long COVID cases are often identified based on one or more persistent symptoms beyond 3 months post-COVID-19.^7^ Long COVID symptoms are both non-specific and common, and present in a substantial proportion of patients without previous SARS-CoV-2 infection.^8^ Therefore, studies using this case-definition without considering severity, functional impact and COVID-19-attribution likely overestimate long COVID prevalence. Most published studies have not assessed long COVID severity or did not compare symptoms between long COVID cases and controls.^9–12^ From a public health perspective, cases severe enough to impact functional status are most relevant and are unlikely to present with the same symptom profile as people without long COVID.

Based on a population survey among healthcare workers (HCWs) in Quebec, Canada, we estimated long COVID risk after first and subsequent COVID-19 infections since the onset of the pandemic, as well as long COVID prevalence and severity in the spring of 2023. We compared long COVID cases and controls on symptom frequency, severity and impact on functional status and identified distinct long COVID phenotypes using clustering analyses.

## METHODS

This study was legally mandated by the National Director of Public Health of Quebec under the Quebec Public Health Act and approved by the CHU de Québec-Université Laval Research Ethics Committee. Electronic or verbal consent was obtained from all participants. The study followed the Strengthening the Reporting of Observational Studies in Epidemiology (STROBE) guideline.

### Study Design and Population

This retrospective cohort study was conducted via an electronic survey among Quebec HCWs from May 16 to June 15, 2023. Eligible participants were HCWs aged ≥18, residing in Quebec and able to communicate in French or English. They had to have been working in the Quebec public healthcare network during the pandemic or registered in one of six professional orders including: physicians, nurses, nursing assistants, respiratory therapists, pharmacists, and midwives.

### Recruitment and Data Collection

#### Online Survey

HCWs received an email invitation to complete a 45-minute self-administered online survey. Data collected included sociodemographic information (sex, age, and race/ethnicity), employment characteristics (occupation and workplace), and COVID-19 history (number of infections, laboratory confirmation, symptoms, severity, and duration of each episode). The survey queried 20 symptoms selected based on reported frequency in the literature and expert opinion.^9,13^ Cases were asked about symptoms related to COVID-19 that lasted ≥12 weeks and were still present at the time of the survey with their severity rated on a three-point scale (mild, moderate, severe). Controls were also queried about symptoms present at the time of survey that had lasted ≥2 weeks. Information on comorbidities was obtained from the Provincial Integrated Chronic Disease Surveillance System of Quebec, updated on March 31, 2021.^14^ Participants reported perceived health status, functional disability due to dyspnea (Modified Medical Research Council Dyspnea [mMRC] Scale), and cognitive impairment on daily life.^15^

#### Telephone Survey

To address possible low participation in the online survey, potentially leading to bias, we conducted a short validation sub-study by phone between May 8 and July 5, 2023, among 7 500 randomly selected non-respondents and 3 000 non-vaccinated HCWs without email access. The telephone survey included 5 questions on COVID-19 history, symptom duration, and persistence at the time of the interview. Consenting participants were invited to complete the online survey.

The recruitment process is described in Supplement_eFigure_1. SARS-CoV-2 variant classification was based on provincial laboratory data on dominant variants at the time of each self-reported infection and reinfection.^16^

#### Long COVID Case and Control Definitions

Long COVID cases were HCWs with self-reported post-COVID-19-attributed symptom(s) lasting ≥12 weeks. Cases were classified as:

a. Prevalent, if any symptom was present at survey time,
b. Recovered, if symptoms had resolved at survey time,
c. Undetermined, if information on symptoms was missing or contradictory.

Long COVID severity was assessed only in prevalent cases, classified as mild (only mild symptoms), moderate (≥1 moderate but no severe symptoms), or severe (≥1 severe symptom).

Controls included participants without self-reported history of COVID-19 (non-COVID controls) and those who had COVID-19 without persistent symptoms at ≥12-week follow-up (COVID controls).

#### Statistical analysis

We compared the sociodemographic and employment characteristics of participants within the target population. Long COVID risk was calculated as: *(Long COVID cases/COVID cases with onset ≥ 12 weeks before survey date).*.

We calculated long COVID risk by number of infections (one to three), likely infecting variant, and severity of the acute COVID-19 episode over the entire study period and specifically for the Omicron period. Reinfection risk was estimated only for participants without long COVID after their first infection. Long COVID prevalence among HCWs was calculated as: *(long COVID cases prevalent at survey date/all participants)*. Binomial 95% confidence intervals (CI) were estimated for both measures.

Using prevalence ratios (PRs), we compared cases and controls on the frequency, severity and number of symptoms. Sensitivity analyses excluded participants who reported the presence of all symptoms listed in the survey, considered unlikely, to assess for potential questionnaire biases.

Cluster analysis of long COVID symptoms was conducted using unsupervised learning with K-means clustering followed by descending hierarchical classification(REF=Hastie).^17^. Cluster numbers were determined using eigenvalues and validated by physicians with clinical expertise in long COVID. The frequency of each cluster was assessed globally and by long COVID severity.

## RESULTS

### Study Population

Among the 397 222 HCWs invited by email to participate in the online survey, 30 237 (7.6%) accessed the survey (Supplement_eFigure_1). Among these, 4 432 declined to participate, 4 287 were excluded primarily due to invalid or incomplete responses (n=3 375) and 21 518 (5.4%) completed the online survey and were included in the analysis.

For the validation study, 53.0% (3 978/7 500) of non-respondents to the email invitation and 36.1% (1 083/3 000) of unvaccinated HCWs answered the five-question telephone survey. Among them, 829 (11.1%) and 149 (5.0%), respectively, thereafter completed the online survey and were included in the final dataset.

Compared to the target population of 397 222 HCWs, online survey participants were comparable in terms of sex, comorbidities and region of residence but were older, received more vaccine doses and were more likely to occupy social services and managerial roles. In contrast, telephone survey participants recruited among non-respondents were broadly representative of the target population (Supplement_eTable_1).

Online survey participants were predominantly female, as generally reflective of the healthcare workforce, self-identified as white, with a median age of 45 years, and worked as nurses, social services professionals, auxiliary staff or managerial and administrative personnel in acute-care hospitals (Table_1). Among 22 496 online participants, 5 434 (24.2%) reported never having had a SARS-CoV-2 infection (non-COVID controls), 13 865 (61.6%) had at least one SARS-CoV-2 infection with onset ≥12 weeks before the survey but without persistent symptoms (COVID controls), and 2 796 (12.4%) had at least one infection with symptoms persisting ≥12 weeks (long COVID cases). Non-COVID controls were more likely to be male, aged ≥60 years and had multiple comorbidities compared to COVID controls across both online and telephone participants. Although COVID-19 attribution was primarily based upon self-reported infection or reinfection, >80% of long-COVID cases and 75% of COVID controls had a documented laboratory-confirmed infection.

### Long COVID Risk and Prevalence

The overall risk of long COVID among HCWs with COVID-19 was similar between online and telephone survey participants: 17.0% (95% CI, 16.3%–17.6%) vs. 15.9% (14.6%–17.2%) (Table_2). Long COVID prevalence at survey time, however, was much lower among telephone survey participants (5.6% [95% CI, 4.9%–6.3%]) than among online survey participants (11.0% [10.5%–11.4%]), with the former likely more representative of the true long COVID prevalence among HCWs.

The cumulative risk of long COVID increased with the number of reported COVID-19 infections, rising from 13.7% (95% CI, 13.1%–14.4%) for a single infection to 37.0% (33.0%–40.9%) for 3 infections in the online survey, and from 11.8% (a single infection) to 29.5% (≥3 infections) in the telephone survey (Figure_1, Table_1, Supplement_eTable_2). However, the risk of long COVID was approximately three times higher after the initial infection (14.8% [95% CI, 14.3%–15.3%]) than after first (5.8% [5.1%–6.6%]) or second (5.3% [3.1%–7.4%]) reinfections.

For each infection, long COVID risk was highest when attributed to the ancestral strain compared to later variants, but still showing the same pattern of higher risk with first infection vs. reinfection for each variant (Figure_1). The risk was also higher among HCWs with severe acute COVID-19, particularly those requiring hospitalization or ambulatory cases reporting ≥3 severe symptoms. Conversely, the risk of long COVID was <5% for HCWs with moderate and mild COVID-19 for both first infection and reinfections. The pattern of highest risk with first infection and with more severe acute COVID-19 persisted in analyses restricted to Omicron cases (Figure_1, Supplement_eTable_3). Omicron infections, responsible for 78.6% of all long COVID cases, were associated with a similar long COVID risk regardless of subvariants (BA.1 to XBB) (Figure_1, Supplement_eFigure_2). Overall, the distribution of long COVID severity was similar for cases occurring after first or second infection, with the highest proportion of severe long COVID cases occurring after a severe acute COVID-19 episode (Supplement_eFigure_3).

### Symptoms at survey time and symptom clustering

Based on self-reported severity, 24.2% (n=515) of long COVID cases were classified as mild, 42.7% (n=907) as moderate, and 33.1% (n=703) as severe (Table_3). Among the 20 symptoms queried, the median number of reported symptoms increased with severity: 2 symptoms (0 moderate or severe) in mild cases, 5 symptoms (2 moderate or severe) in moderate cases and 11 symptoms (7 moderate or severe) in severe cases. In contrast, 60.8% of COVID controls and 51.0% of non-COVID controls reported no symptoms at survey time, with a median of 3 (1 moderate or severe) symptoms among those experiencing any.

The most frequently reported symptoms among long COVID cases were fatigue, shortness of breath and cognitive symptoms (Supplement_eFigure_4). Each symptom was reported at least twice as often by participants with long COVID compared to COVID controls, with PRs ranging from 2.1 to 5.1, highest for shortness of breath, smell/taste disturbance, post-exertional malaise (PEM), and brain fog (Figure_2). Severe symptoms were reported 5 to 22 times more often by long COVID cases than by COVID controls, except for fever, cough, insomnia, anxiety, and depression (2.7 to 4.5 times). In sensitivity analyses excluding cases and controls reporting all symptoms, differences between cases and controls were not diminished (Supplement_eTable_4. Supplement_eFigure_5, Supplement_eFigure_6).

Clustering analyses identified seven distinct symptom clusters, explaining 75.9% of total variance (Supplement_eFigure_7), that are detailed by severity of long COVID in Supplement_eFigure_8. The most common clusters globally and among severe cases included systemic, neurocognitive, pulmonary and mood-related symptoms. Overall, 19.3% of cases exhibited symptoms only from a single cluster; 42.8% in mild, 16.5% in moderate and 5.6% in severe cases. Smell/taste impairment was the most frequent cluster reported in isolation.

Among 1 329 prevalent long COVID cases reporting a single COVID-19 episode, the mean duration of long COVID symptoms was 15 months, with 693 (52.1%) experiencing symptoms for ≥1 year, and 83 (6.2%) reporting symptoms lasting ≥3 years (Supplement_eTable_5). Recovered long COVID cases had a mean symptom duration of seven months, with 93 (61.6%) recovering within six months.

### Functional limitations

Long COVID cases more often than COVID and non-COVID controls reported poor self-rated health (29.5% vs. 5.5% and 9.4%), moderate or severe effort performance limitations (23.0% vs. 3.4% and 5.6%), and cognitive impairments affecting daily life (e.g., 18.6% often/very often losing necessary items vs. about 6% among controls). These limitations were significantly more pronounced in severe long COVID cases (Table_3).

## DISCUSSION

In this large population-based study of Quebec HCWs, conducted in the spring of 2023, the estimated overall risk of long COVID was 16-17%. This risk was highest following infection by the ancestral strain and lowest after Omicron infection that nevertheless caused most long COVID cases given its widespread transmission. Long COVID risk was higher after first infection compared to reinfection, whether by all variants considered together (14% vs. 5-6%) or Omicron only (13% vs. 3-5%). As of May 2023, the estimated long COVID prevalence among Quebec HCWs was 5.6%. Symptom number, severity and duration, clusters of symptoms and functional profiles were substantially different between moderate or severe long COVID cases vs. controls.

The range of long COVID risk estimates in the published literature reflects differences in study periods, populations, case-definitions and case ascertainment approaches.^4,5,18–20^ Our 16-17% overall risk of long COVID is comparable to estimates in various North American and UK cohorts. For instance the risk of long COVID was 22% in 14 cohorts followed from 2020 to 2023 in the United States,^21^ 15% among American adults with test-confirmed COVID-19,^22^ 19% among Canadian adults with self-reported COVID-19,^23^ and 8-17% in 10 community-based UK cohorts with varying long COVID definitions.^24^ The higher risk of long COVID estimated in our study following infection by the ancestral strain, and lowest risk after Omicron infection corroborates previous observations.^19–22,25^ The very high vaccination uptake in our cohort of HCWs may have contributed to the lower risk after Omicron infections.

Whereas some previous studies have reported lower long COVID risk with repeat infection,^26,27^ our study indicates that long COVID risk is roughly two thirds lower following reinfection compared to first infection. This may be partly related to greater host-specific resistance among individuals who did not have long COVID following their first episode. The cumulative risk of long COVID during the study period increased with the number of infections, a trend also observed elsewhere.^23,26^ Even if the per episode risk is highest with first infection, the higher cumulative risk for individuals with repeated infections reflects ongoing accumulation of the persisting risk with reinfections.

The prevalence of long COVID depends upon the level of SARS-CoV-2 transmission, its associated long COVID risk, the duration of long COVID condition and the duration of follow-up. At the time of our May 2023 survey, 5.6% of telephone participants and 11.0% of online participants reported symptoms consistent with long COVID. Despite the smaller telephone survey sample, their prevalence likely better represents the true HCW population estimate given the higher response rate and representativeness. A nationwide Canadian survey estimated that 7% of Canadian adults had long COVID in 2022.^23^ According to the U.S. National Health Interview Survey of 2022, 6.9% of American adults ever had long COVID, 3.4% in 2022,^28^ while the Household Pulse Survey found that 17.6% ever had long COVID, 6.9% in 2024.^1^ In the UK, 3% of the population was estimated to have long COVID in March 2023.^28^

In the absence of diagnostic biomarkers, long COVID ascertainment is based on non-specific signs or symptoms.^29^ The most prevalent symptoms in our study were fatigue, shortness of breath, neurocognitive symptoms and PEM, arranged within systemic, neurocognitive and respiratory clusters, which aligns with other population-based and clinical studies.^1,7,9,30–36^ Previous studies comparing symptoms (of any severity) reported by COVID-19 infected and uninfected participants found a high prevalence of core long COVID symptoms also among uninfected individuals.^8,36–39^ Prevalence ratios comparing infected vs. uninfected participants were relatively low (at 0.8 to 3) even for more specific symptoms like smell/taste disturbance. When we compared prevalent long COVID cases and COVID controls, PRs were high at 2-5 for symptoms of any severity and much higher for severe or multiple (≥5) symptoms. This suggests that future studies assessing risk factors of long COVID may benefit from using both the current sensitive but less specific, as well as more specific but less sensitive case-definitions, the latter requiring several symptoms and/or a moderate-severe experience. If one uses the “standard” case definition, it seems important to not only document the symptoms but also to annotate those specifically attributed by patients to a COVID-19 episode.

Long COVID patients report fatigue, shortness of breath and cognitive symptoms as the most debilitating symptoms.^9^ We found that cases overall were 2-3 times, and severe cases 4-10 times, more likely than controls to report dyspnea-associated and cognitive impairments overall. Post-COVID-19 limitations in physical performance effort have been described after hospitalized and ambulatory COVID-19 episodes.^40–42^ Among patients from long COVID clinics, 36% and 40%, respectively, reported moderate and severe exercise limitations compared to their pre-COVID-19 performance, indicating drastic decrease in the frequency of moderate or intense physical activity.^43,44^ Cognitive impairments associated with long COVID have been described elsewhere, but their association with long COVID severity has not been previously documented.^33,43,45^

### Strengths and limitations

Our large and systematic HCW-based study expands understanding of the evolving long COVID risk during the first three years of the pandemic. For the first time, we describe the complex relationships of long COVID risk and severity with SARS-CoV-2 variants, acute COVID-19 severity and first infection vs. reinfections. Moreover, we show that a survey-based classification of long COVID severity is associated with the specificity of the case-definition and with functional impact.

Our main limitation was the low response rate, which likely overestimated long COVID prevalence. Nevertheless, our validation telephone sub-study with a higher response rate from a representative sample of the target population found similar results for long COVID risk after first infection vs. reinfection, as well as cumulative risk with number of infections, reassuring that other associations assessed through online participants were likely not biased. The retrospective observational design might have led to some recall bias, potentially affecting precision for the number and exact timing of COVID-19 episodes, duration of symptoms (based on infection date) and identification of recovered long COVID cases (based on symptom start and end dates). Our study sample consisted mostly of middle-aged white female HCWs, reflective of the healthcare workforce in Quebec, which may limit generalizability to other populations.

## Conclusion

Long COVID is a common and disabling condition among HCWs. Societal and healthcare burden remains important and will continue to accrue given ongoing SARS-CoV-2 transmission and long COVID risk with reinfections.

## Data Availability

Underlying data cannot be shared publicly by authors, because it belongs to the Ministere de la sante et des services sociaux du Quebec and data access to researchers was given under the legal mandate of the Quebec National Director of Public Health. A request for data should be addressed to the research access centre designated by Quebec Government in accordance with the act respecting health and social services information (https://statistique.quebec.ca/en/institut/services-for-researchers/data).

## Acknowledgments

We thank Stéphanie Grenier, Josiane Rivard and Sandrine Hegg-Deloye (CHU de Québec-Université Laval Research Center) for their work preparing and administering the online and telephone surveys, Meghan Kanou (Montreal University) for her contribution in the literature review, Maria Tran (Institut National de Santé Publique du Québec) for her contribution with some statistical analyses. Finally, we thank all the participants enrolled in the Quebec healthcare worker cohort.

---

**Table 1.** Characteristics of the study population.

| Characteristic | Online survey |  |  | Telephone survey<sup>a</sup> |  |  |
| --- | --- | --- | --- | --- | --- | --- |
|  | Long COVID Cases | COVID Controls<sup>b</sup> | Non-COVID Controls | Long COVID Cases | COVID Controls | Non-COVID Controls |
| **N** | 2796 | 13865 | 5434 | 596 | 3283 | 1116 |
| **Sex** |  |  |  |  |  |  |
| Female | 2374 (84.9) | 11320 (81.6) | 4141 (76.2) | 495 (83.1) | 2638 (80.4) | 809 (72.5) |
| Male | 422 (15.1) | 2545 (18.4) | 1293 (23.8) | 101 (16.9) | 645 (19.6) | 307 (27.5) |
| **Age, years** |  |  |  |  |  |  |
| Median (IQR) | 45.9<br>(38.2-54.1) | 43.8<br>(35.2-53.7) | 50.3<br>(39.5-59.6) | 43.2<br>(33.4-53.3) | 39.4<br>(30.4-50.7) | 46.0<br>(34.3-57.4) |
| 18-39 | 827 (29.6) | 5318 (38.4) | 1415 (26.0) | 243 (40.8) | 1686 (51.4) | 415 (37.2) |
| 40-49 | 951 (34.0) | 4047 (29.2) | 1266 (23.3) | 162 (27.2) | 737 (22.4) | 245 (22.0) |
| 50-59 | 709 (25.4) | 2821 (20.3) | 1457 (26.8) | 128 (21.5) | 555 (16.9) | 244 (21.9) |
| ≥60 | 309 (11.1) | 1679 (12.1) | 1296 (23.8) | 63 (10.6) | 305 (9.3) | 212 (19.0) |
| **Race / ethnicity** |  |  |  |  |  |  |
| White | 2296 (82.1) | 12049 (86.9) | 4282 (78.8) | NA | NA | NA |
| Black | 157 (5.6) | 538 (3.9) | 501 (9.2) | NA | NA | NA |
| Hispanic | 78 (2.8) | 259 (1.9) | 79 (1.5) | NA | NA | NA |
| Arab | 61 (2.2) | 222 (1.6) | 156 (2.9) | NA | NA | NA |
| Asian | 63 (2.3) | 362 (2.6) | 168 (3.1) | NA | NA | NA |
| Other / no response | 141 (5.0) | 435 (3.1) | 248 (4.6) | NA | NA | NA |
| **Born in Canada** | 2299 (82.2) | 11888 (85.7) | 4254 (78.3) | NA | NA | NA |
| **Chronic conditions** |  |  |  |  |  |  |
| None | 1398 (50.0) | 8262 (59.6) | 3046 (56.1) | 311 (52.2) | 1960 (59.7) | 652 (58.4) |
| At least two<sup>c</sup> | 593 (21.2) | 2048 (14.8) | 956 (17.6) | 111 (18.6) | 461 (14.0) | 163 (14.6) |
| Chronic heart disease | 152 (5.4) | 539 (3.9) | 259 (4.8) | 27 (4.5) | 126 (3.8) | 51 (4.6) |
| Chronic lung disease | 340 (12.2) | 1056 (7.6) | 430 (7.9) | 61 (10.2) | 233 (7.1) | 79 (7.1) |
| History of depression | 512 (18.3) | 1771 (12.8) | 653 (12.2) | 101 (16.9) | 484 (14.7) | 141 (12.6) |
| Obesity | 199 (7.1) | 684 (4.9) | 269 (5.0) | 40 (6.7) | 122 (3.7) | 40 (3.6) |
| Hypertension | 248 (8.9) | 987 (7.1) | 625 (11.5) | 51 (8.6) | 191 (5.8) | 93 (8.3) |
| Diabetes | 125 (4.5) | 436 (3.1) | 292 (5.4) | 31 (5.2) | 92 (2.8) | 49 (4.4) |
| **Occupation** |  |  |  |  |  |  |
| Physician | 93 (3.3) | 775 (5.6) | 281 (5.2) | 9 (1.5) | 127 (3.9) | 48 (4.3) |
| Nursing and respiratory care | 838 (30.0) | 3728 (26.9) | 1291 (23.8) | 177 (29.7) | 925 (28.2) | 230 (20.6) |
| Paratechnical personnel and auxiliary services | 592 (21.2) | 1938 (14.0) | 1134 (20.9) | 193 (32.4) | 851 (25.9) | 392 (35.1) |
| Health and social services technicians and professionals | 651 (23.3) | 3977 (28.7) | 1167 (21.5) | 87 (14.6) | 645 (19.6) | 168 (15.1) |
| Administration / manager | 558 (20.0) | 2918 (21.1) | 1390 (25.6) | 116 (19.5) | 632 (19.3) | 251 (22.5) |
| Other / Unknown | 64 (2.3) | 529 (3.8) | 171 (3.2) | 14 (2.3) | 103 (3.1) | 27 (2.4) |
| **Workplace** |  |  |  |  |  |  |
| Acute-care hospital | 1180 (42.2) | 6160 (44.4) | 2372 (43.7) | NA | NA | NA |
| Nursing home | 520 (18.6) | 1607 (11.6) | 771 (14.2) | NA | NA | NA |
| Clinics / health centers | 403 (14.4) | 2257 (16.3) | 734 (13.5) | NA | NA | NA |
| Other | 693 (24.8) | 3841 (27.7) | 1557 (28.7) | NA | NA | NA |
| **COVID-19 vaccination<sup>d</sup>** |  |  |  |  |  |  |
| Unvaccinated | 17 (0.6) | 117 (0.8) | 37 (0.7) | 102 (17.1) | 670 (20.4) | 292 (23.2) |
| 1 dose | 41 (1.5) | 108 (0.8) | 8 (0.1) | 17 (2.9) | 32 (1.0) | 2 (0.2) |
| 2 doses | 539 (19.3) | 1936 (14.0) | 680 (12.5) | 150 (25.2) | 681 (20.7) | 203 (18.2) |
| 3 or more doses | 2199 (78.6) | 11704 (84.4) | 4709 (86.7) | 327 (54.9) | 1900 (57.9) | 619 (55.5) |
| **COVID-19 episodes** |  |  |  |  |  |  |
| 0 | NA | NA | 5434 (100.0) | NA | NA | 1116 (100.0) |
| 1 | 1687 (60.3) | 10586 (76.4) | NA | 297 (49.8) | 2225 (67.8) | NA |
| 2 | 865 (30.9) | 2890 (20.8) | NA | 231 (38.8) | 885 (27.0) | NA |
| 3 or more | 244 (8.7) | 244 (2.8) | NA | 67 (11.2) | 160 (4.9) | NA |
| Self-reported NAAT confirmation (one or several episodes) | 2331 (83.4) | 10424 (75.2) | NA | NA | NA | NA |

<sup>a</sup> Telephone survey participants were not asked about their race / ethnicity, their native country, their workplace; 14 participants did not know the number of COVID-19 episodes.
<sup>b</sup> Participants reporting a single COVID-19 episode occurring <84 days before survey completion were not included.
<sup>c</sup> At least two chronic conditions among the following: alcohol abuse, anaemia, cancer, cardiovascular disease, chronic respiratory disease, coagulopathy, diabetes, drug abuse, fluid and electrolyte disorders, history of depression, hypertension, hypothyroidism, kidney disease, liver disease, neurological disorder, obesity, psychosis, ulcer, paralysis, weight loss.
<sup>d</sup> Number of COVID-19 vaccine doses received at the time of survey.
Abbreviations: IQR, interquartile range; NA, not available / not applicable; NAAT, nucleic acid amplification test.

**Table 2.** Risk and prevalence of long COVID by type of survey and type of recruitment.

| Recruitment | Online survey |  |  | Telephone survey |  |
| --- | --- | --- | --- | --- | --- |
|  | By email | By telephone among non respondents | By telephone among non vaccinated | By telephone among non respondents | By telephone among non vaccinated |
| **N** |  |  |  |  |  |
| Number of participants | 21518 | 829 | 149 | 3978 | 1083 |
| HCW without COVID-19 episodes, n (%) | 5199 (24.2) | 204 (24.6) | 31 (20.8) | 825 (20.7) | 291 (26.9) |
| HCW with COVID-19 episode(s), n (%) | 16319 (75.8) | 625 (75.4) | 118 (79.2) | 3153 (79.3) | 792 (73.1) |
| HCW with COVID-19 episode(s) ≥12 weeks before survey, n (%) | 15926 (74.0) | 618 (74.5) | 117 (78.5) | 3105 (78.1) | 774 (71.5) |
| Long COVID cases, n (%) | 2700 (12.5) | 81 (9.8) | 15 (10.1) | 494 (12.4) | 102 (9.4) |
| Prevalent long COVID cases, n (%) | 2360 (11.0) | 66 (8.0) | 11 (7.4) | 221 (5.6) | 56 (5.2) |
| Recovered long COVID cases, n (%) | 281 (1.3) | 13 (1.6) | 2 (1.3) | 273 (6.9) | 46 (4.3) |
| Undetermined long COVID cases, n (%) | 59 (0.3) | 2 (0.2) | 2 (1.3) | NA | NA |
| **Long COVID risk among HCWs with COVID-19 episode(s), % (95% CI) <sup>a</sup>** | 17.0<br>(16.3 – 17.6) | 13.1<br>(10.4 – 16.2) | 12.8<br>(7.4 – 20.6) | 15.9<br>(14.6 – 17.2) | 13.2<br>(10.8 – 15.6) |
| **Long COVID prevalence among HCWs with COVID-19 episode(s), % (95% CI) <sup>b</sup>** | 14.8<br>(14.3 – 15.4) | 10.7<br>(8.2 – 13.1) | 9.4<br>(4.1 – 14.7) | 7.1<br>(6.2 – 8.0) | 7.2<br>(5.4 – 9.1) |
| **Long COVID prevalence among participant HCWs, % (95% CI) <sup>c</sup>** | 11.0<br>(10.5 – 11.4) | 8.0<br>(6.2 – 10.0) | 7.4<br>(3.8 – 12.8) | 5.6<br>(4.9 – 6.3) | 5.3<br>(3.9 – 6.6) |

<sup>a</sup> Risk among HCWs with COVID-19 episode(s): Long COVID cases (prevalent, recovered, and undetermined cases) divided by all HCWs with COVID-19 episode(s) with onset at least 12 weeks before survey's date
<sup>b</sup> Prevalence among HCWs with COVID-19 episode(s): Prevalent long COVID cases divided by all HCWs with COVID-19 episode(s) with onset at least 12 weeks before survey's date
<sup>c</sup> Prevalence among all participant HCWs: Prevalent long COVID cases divided by all HCWs
Abbreviations: CI, confidence interval; HCWs, healthcare workers.

**Table 3.**
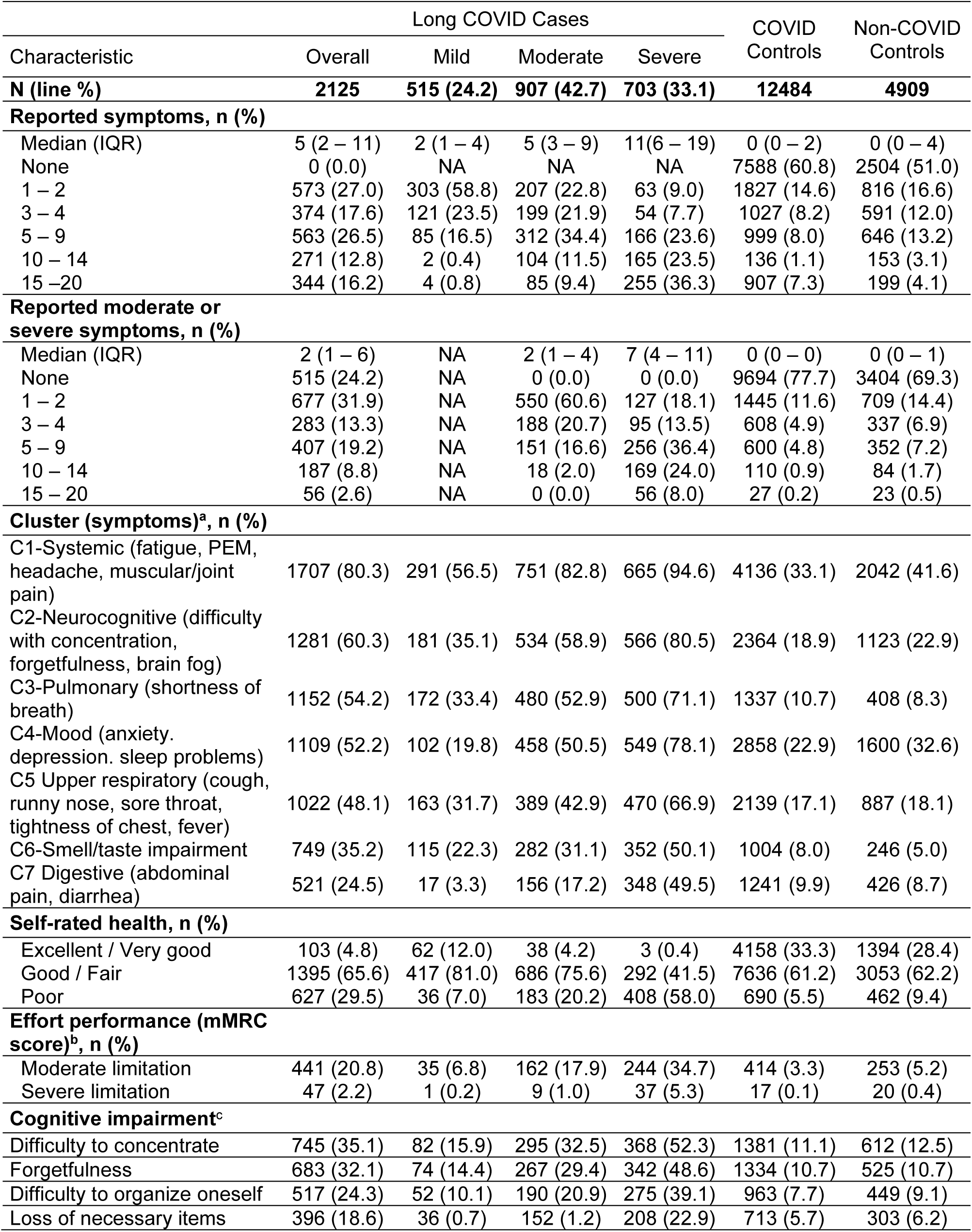
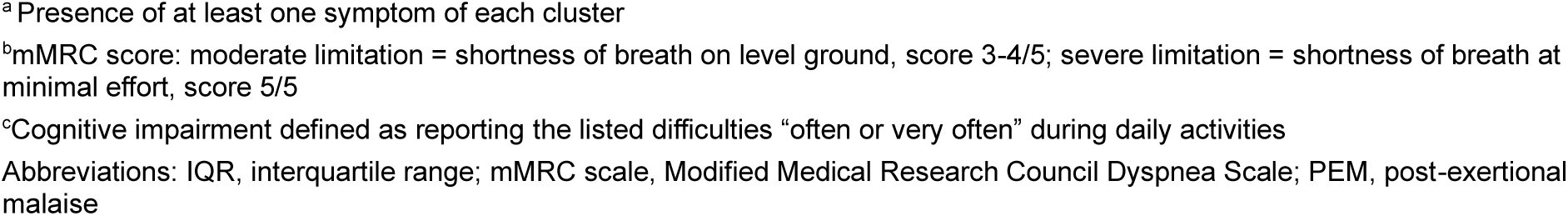
Disease severity, self-rated health and physical and cognitive impairment during daily activities.

**Figure 1.**
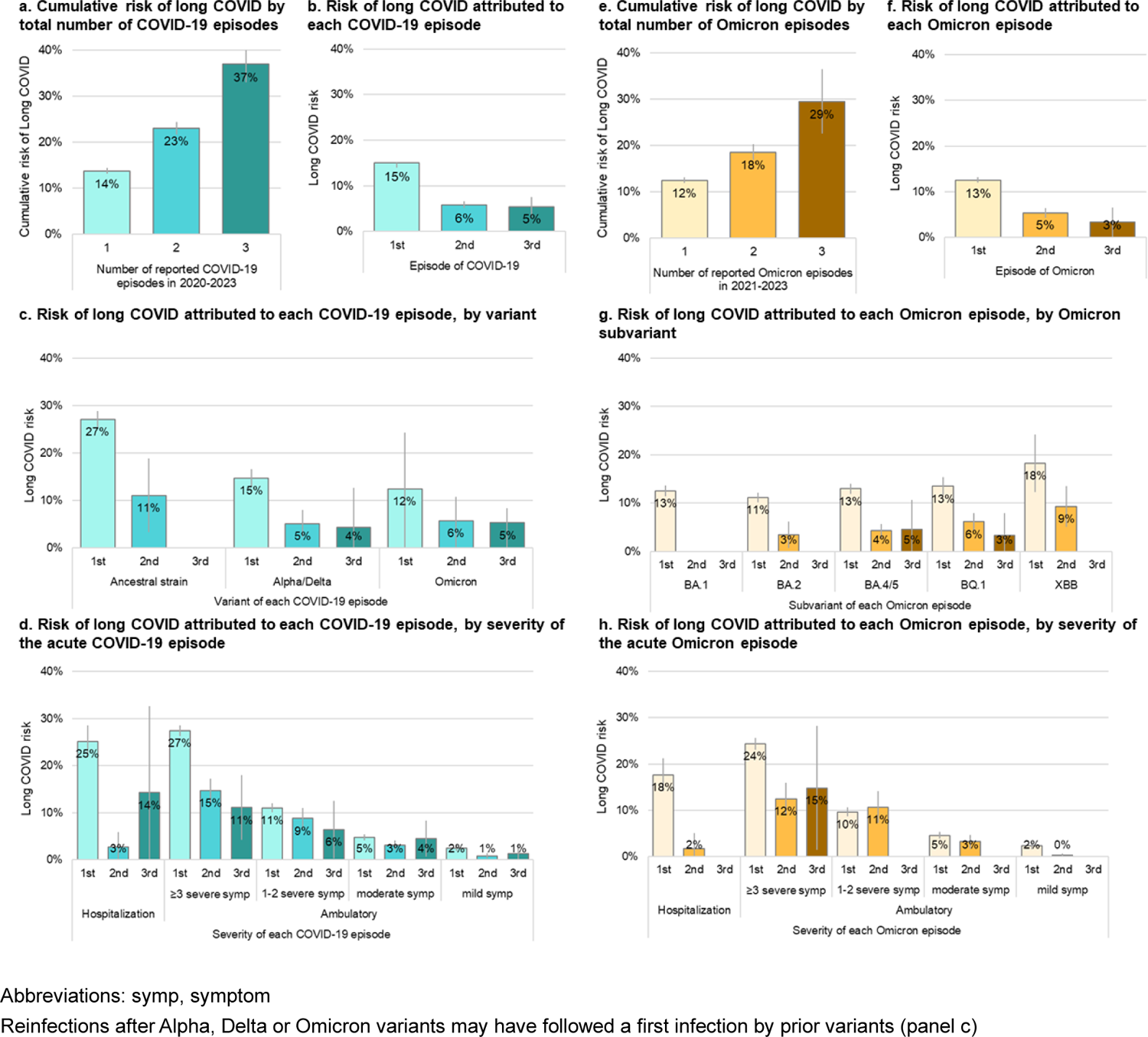
Cumulative long COVID risk by total number of COVID-19 episodes and long COVID risk with initial and subsequent COVID-19 infections by variant and severity of the acute COVID-19 episode

**Figure 2.**
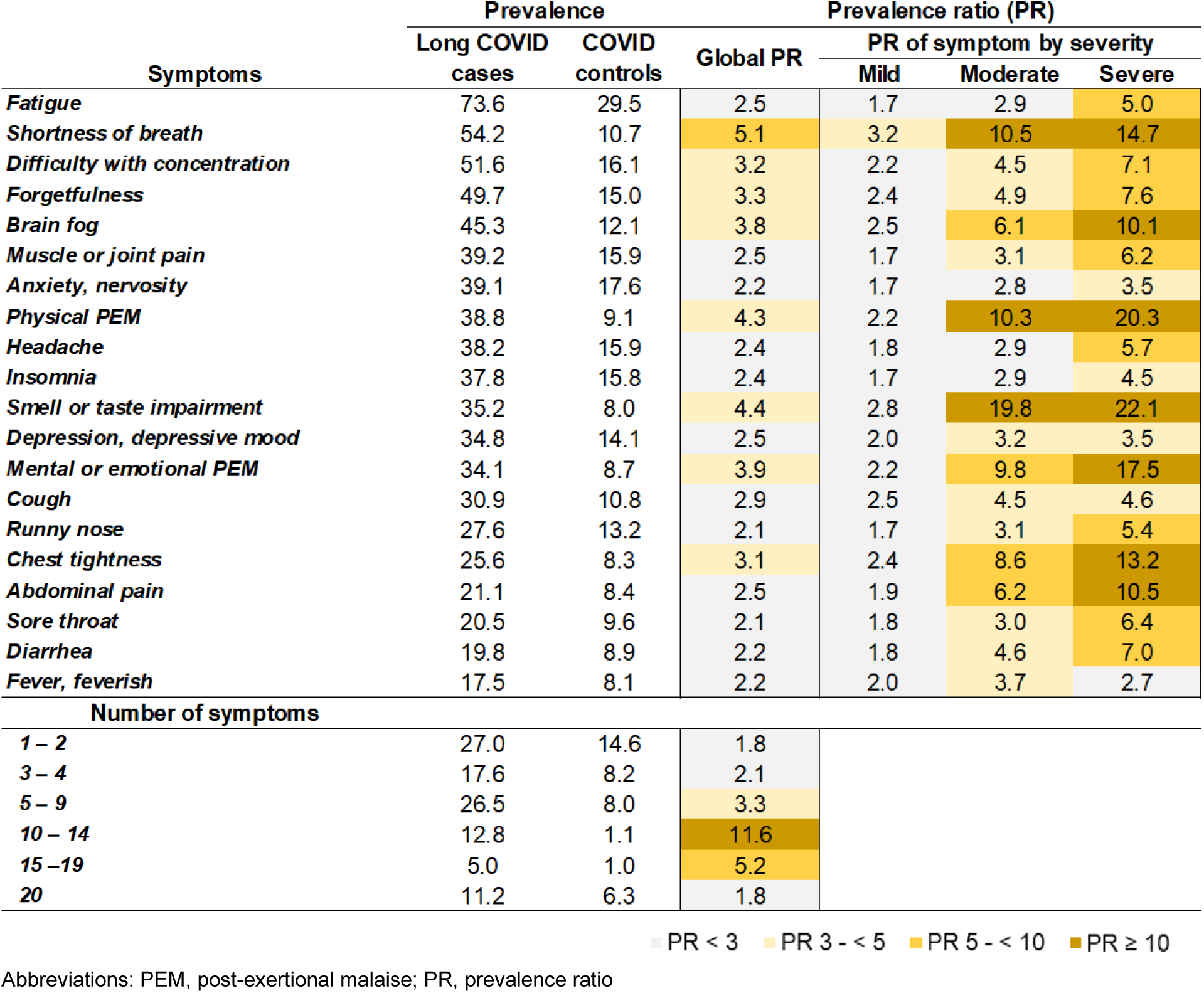
Prevalence ratios of symptoms, by presence and severity, and of number of symptoms comparing long COVID cases and COVID controls

