## Supplemental material for "Long COVID risk and severity after COVID-19 infections and reinfections in Quebec healthcare workers"

eTable 3. Long COVID cases and total Omicron cases by number of infections and episode of attribution of long COVID, among HCWs who had only Omicron infections 12

eTable 4. Disease severity, self-rated health and physical and cognitive impairment during daily activities, after exclusion of participants reporting all symptoms ..... 13

eTable 5. Symptom duration of long COVID cases after first (and single) COVID-19 episode ..... 15

### eFigure 1. Population flowchart

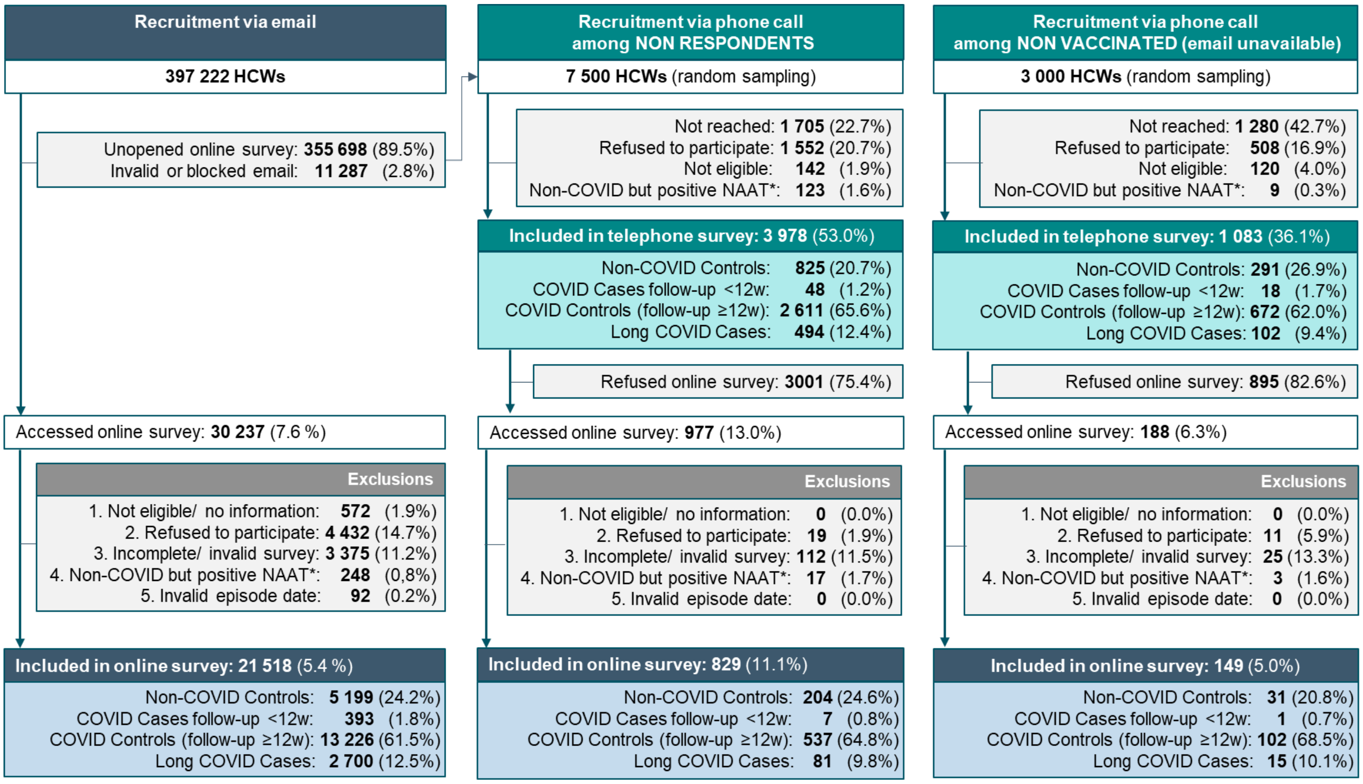

* Exclusion of participants who declared not having had any COVID-19 episode, but the provincial laboratory registry indicated a positive SARS-CoV-2 nucleic acid amplification test

Abbreviations: HCWs, healthcare workers; NAAT, nucleic acid amplification test; w, week

**eFigure 2. Distribution of prevalent long COVID cases after first (and single) episode by long COVID severity and dominant variant at the time of infection**

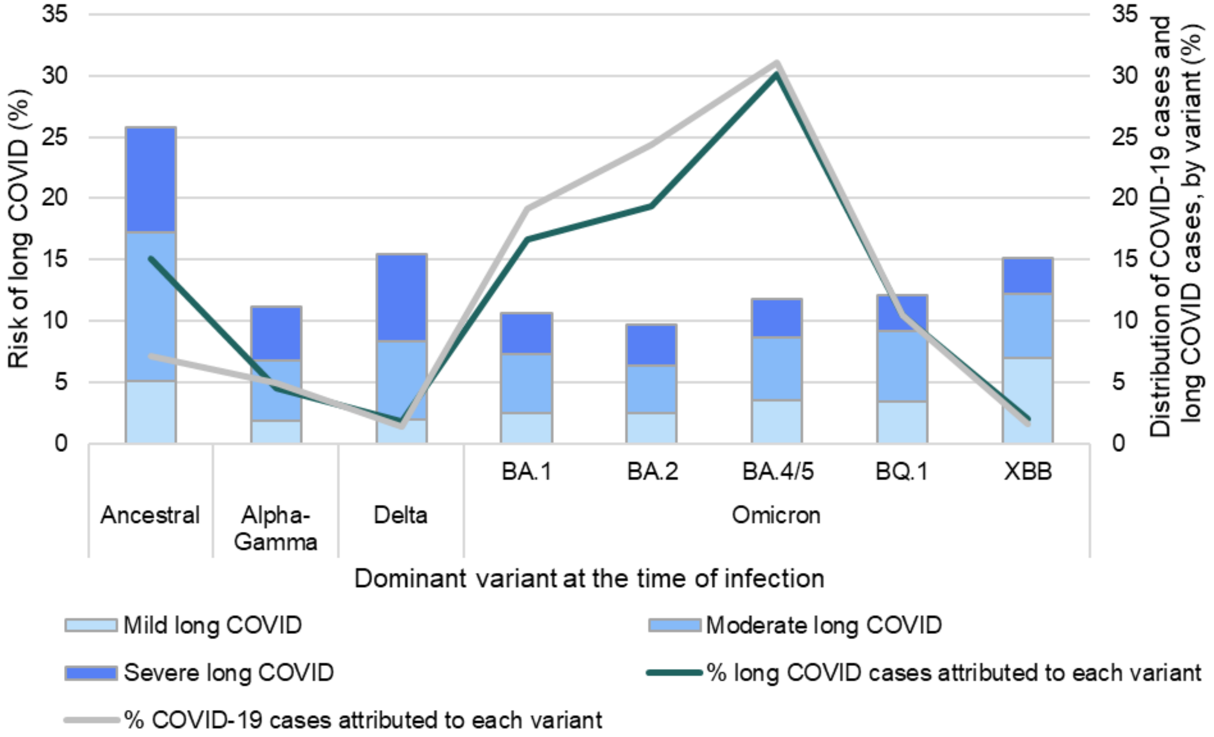

**eFigure 3. Distribution of severity of long COVID after first or second COVID-19 infection by variant and severity of the acute COVID-19 episode**

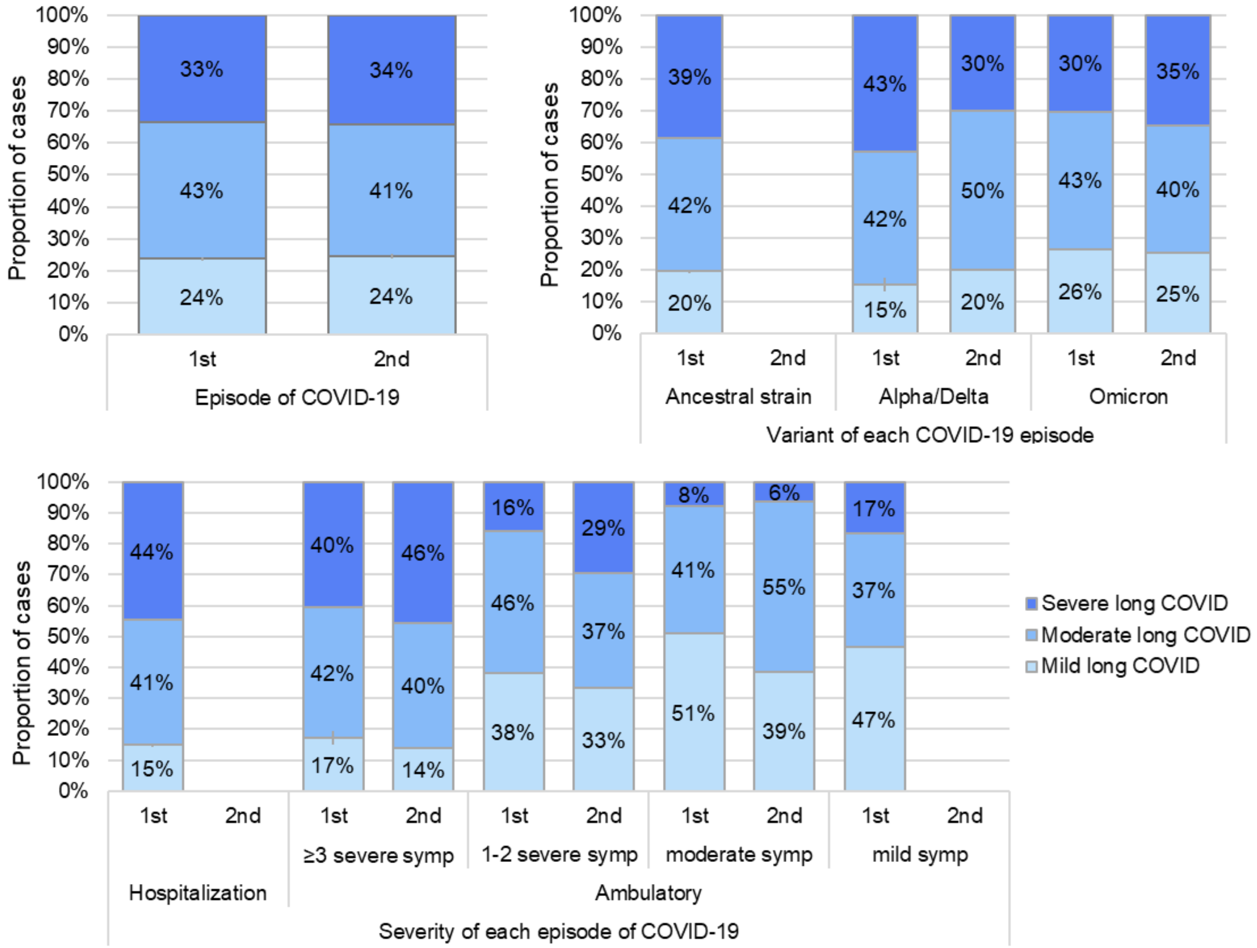

Abbreviations: symp, symptom

### eFigure 4. Most frequent symptoms at the time of the survey reported by prevalent long COVID cases and by COVID and non-COVID controls

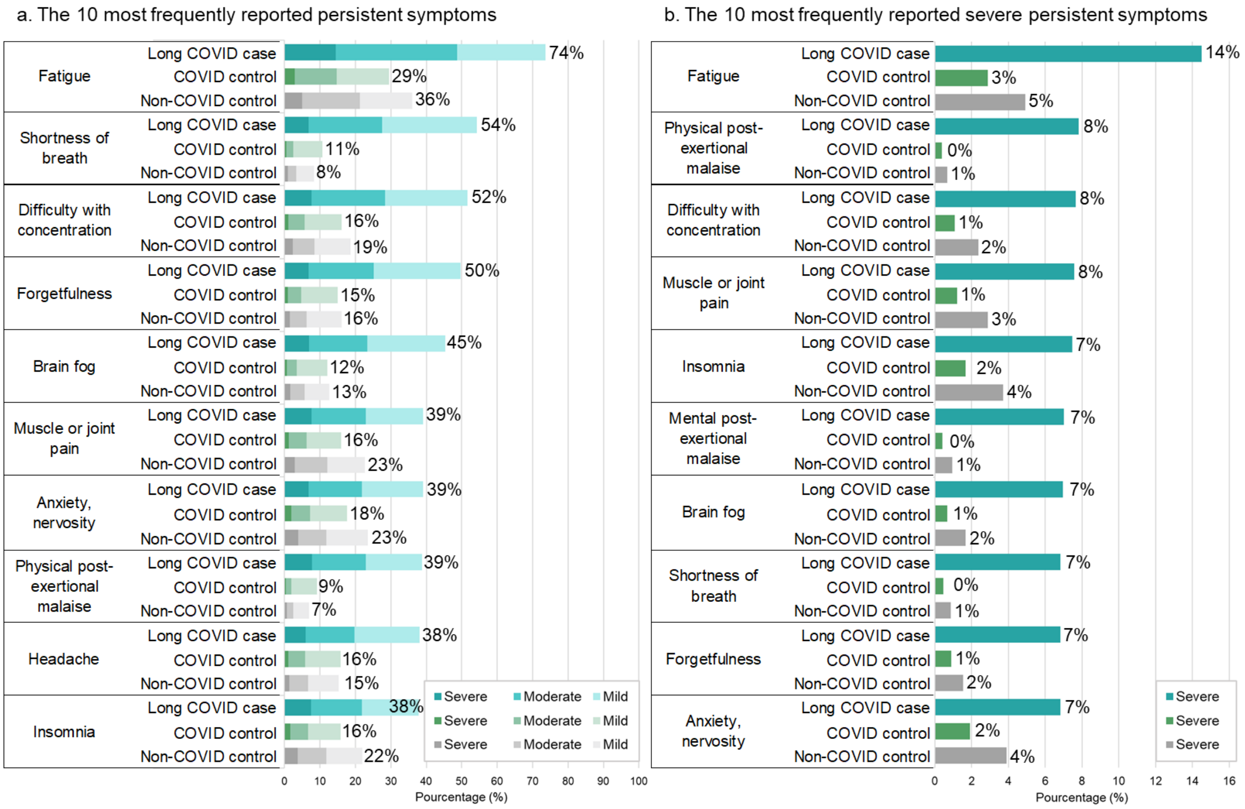

Note: Percentages in the figure correspond to the number of participants who reported each symptom (a) or each severe symptom (b), divided by the total number of participants in each subpopulation

**eFigure 5. Most frequently persistent symptoms reported by prevalent long COVID cases and corresponding frequency of reporting by COVID and non-COVID controls, after exclusion of participants reporting all symptoms**

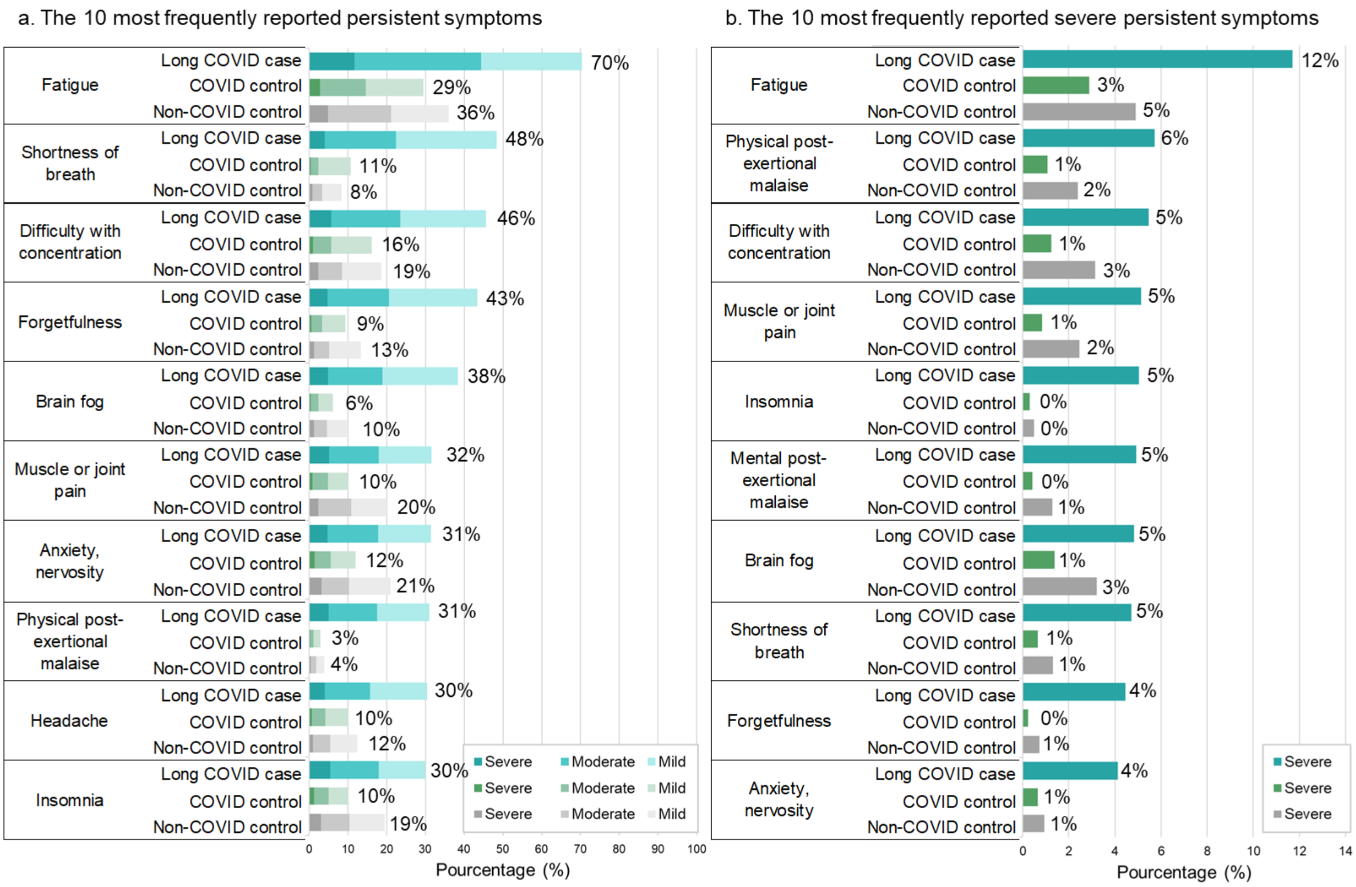

Note: Percentages in the figure correspond to the number of participants who reported each symptom (a) or each severe symptom (b), divided by the total number of participants in each subpopulation after exclusion of participants reporting all symptoms (237 (11%) long COVID cases, 787 (6%) COVID controls and 154 (3%) non-COVID controls)

### eFigure 6. Symptom prevalence ratios of long COVID cases compared with COVID controls, after exclusion of participants reporting all symptoms

| Symptoms | Prevalence | | Prevalence ratio (PR) | | | |
|---|---|---|---|---|---|---|
| | Long COVID cases | COVID controls | Global PR | PR of symptom by severity: Mild | Moderate | Severe |
| *Fatigue* | 70.3 | 24.7 | 2.8 | 2.0 | 3.3 | 5.3 |
| *Shortness of breath* | 48.5 | 4.7 | 10.3 | 8.3 | 14.6 | 14.0 |
| *Difficulty with concentration* | 45.6 | 10.5 | 4.3 | 3.5 | 5.2 | 7.8 |
| *Forgetfulness* | 43.4 | 9.3 | 4.7 | 3.8 | 5.9 | 7.3 |
| *Brain fog* | 38.5 | 6.2 | 6.2 | 5.2 | 7.2 | 11.5 |
| *Muscle or joint pain* | 31.5 | 10.3 | 3.1 | 2.5 | 3.2 | 6.1 |
| *Anxiety, nervosity* | 31.5 | 12.0 | 2.6 | 2.1 | 3.1 | 3.5 |
| *Physical PEM* | 31.1 | 3.0 | 10.4 | 7.1 | 15.6 | 16.8 |
| *Headache* | 30.4 | 10.2 | 3.0 | 2.4 | 3.3 | 6.2 |
| *Insomnia* | 30.0 | 10.1 | 3.0 | 2.3 | 3.4 | 4.4 |
| *Smell or taste impairment* | 27.1 | 1.9 | 14.6 | 9.7 | 31.2 | 29.4 |
| *Depression, depressive mood* | 26.6 | 8.3 | 3.2 | 2.9 | 3.5 | 3.6 |
| *Mental or emotional PEM* | 25.8 | 2.6 | 10.1 | 7.2 | 14.0 | 20.0 |
| *Cough* | 22.2 | 4.8 | 4.7 | 4.8 | 4.5 | 3.3 |
| *Runny nose* | 18.5 | 7.3 | 2.5 | 2.1 | 3.3 | 4.6 |
| *Chest tightness* | 16.3 | 2.1 | 7.7 | 6.9 | 10.6 | 8.9 |
| *Abdominal pain* | 11.2 | 2.3 | 4.9 | 4.0 | 7.1 | 9.5 |
| *Sore throat* | 10.5 | 3.6 | 3.0 | 2.8 | 3.0 | 4.5 |
| *Diarrhea* | 9.7 | 2.8 | 3.5 | 2.8 | 5.7 | 4.5 |
| *Fever, feverish* | 7.2 | 1.9 | 3.8 | 4.0 | 2.9 | 3.1 |
| **Number of symptoms** | | | | | | |
| *1 – 2* | 30.3 | 15.9 | 1.8 | | | |
| *3 – 4* | 19.8 | 8.8 | 2.1 | | | |
| *5 – 9* | 29.8 | 8.5 | 3.3 | | | |
| *10 – 14* | 14.4 | 1.2 | 11,6 | | | |
| *15 –19* | 5.7 | 1.0 | 2,2 | | | |

PR < 3 PR 3 - < 5 PR 5 - < 10 PR ≥ 10

Abbreviations: PEM, post-exertional malaise; PR, prevalence ratio

### eFigure 7. Dendrogram representing hierarchical clustering of long COVID symptoms

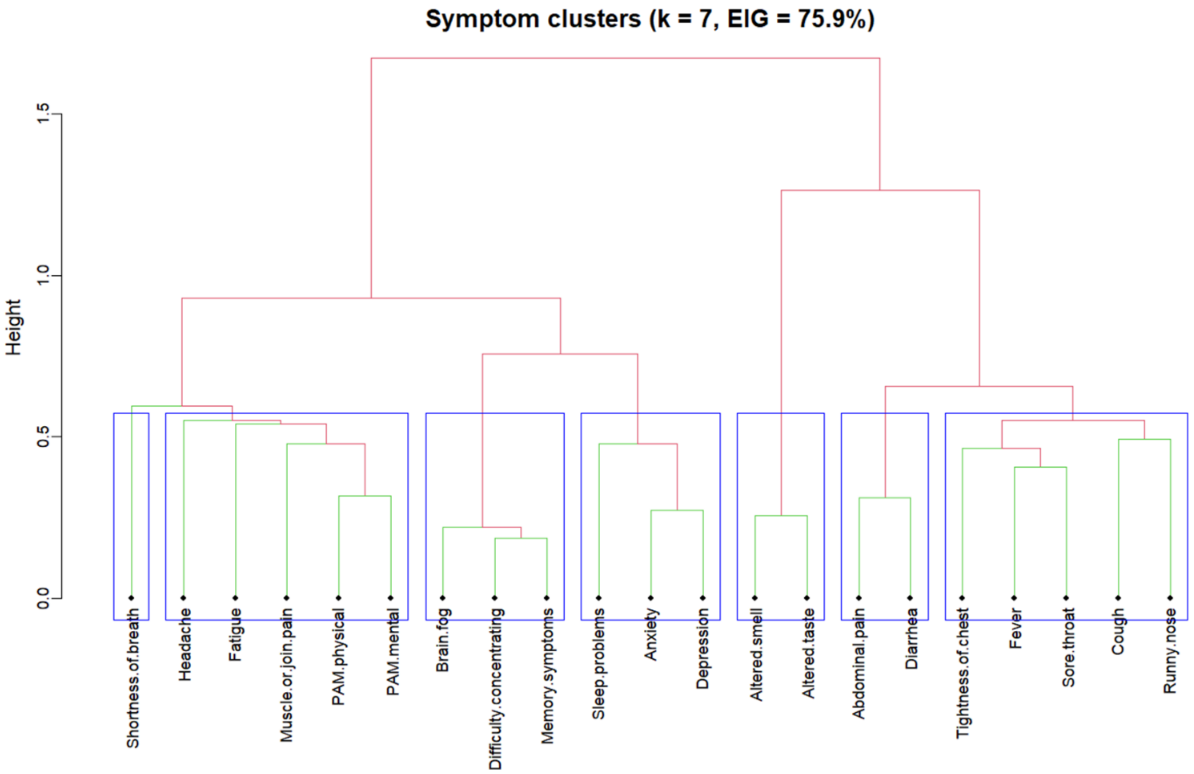

Abbreviations: EIG, eigenvalues of variance; PEM, post-exertional malaise; k, number of clusters

Note: A descending hierarchical clustering analysis, starting from a single grouping of all symptoms to a progressive subdivision leads to seven distinct symptom clusters, explaining 75.9% of the total variance.

### eFigure 8. Cluster distribution of symptoms and proportion of cases affected by each cluster (alone or combined), by long COVID severity

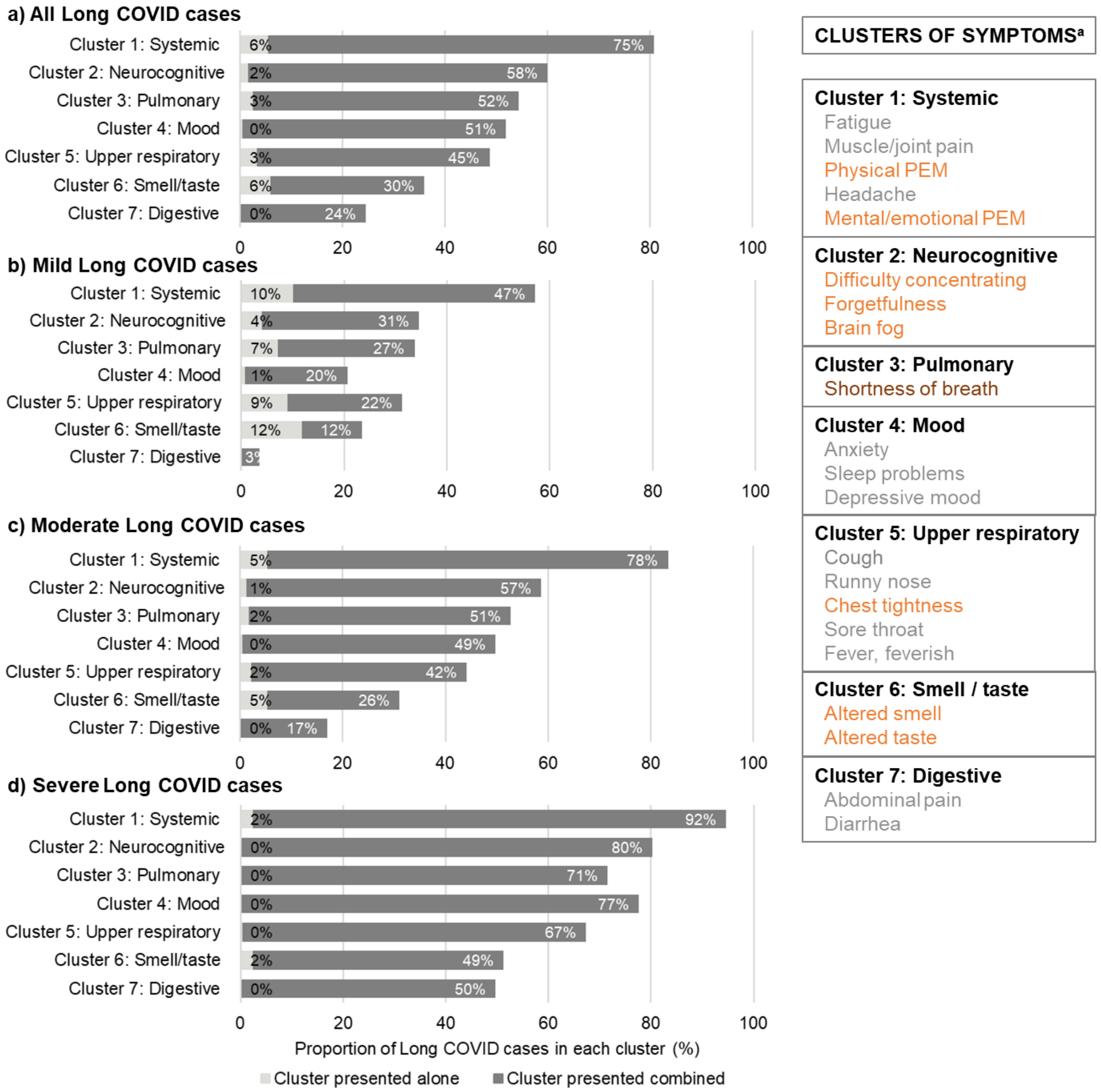

[a]Clusters of symptoms and symptoms within clusters presented by order of reported frequency, with colors indicating the relative prevalence compared with COVID controls (prevalence ratio < 3 (grey), 3 -< 5 (orange), ≥ 5 (brown))

Abbreviations: PEM, post-exertional malaise.

### eTable 1. Characteristics of targeted population and of participants in the online and telephone survey

| Population | Target population of HCWs invited by email | | Participants to online survey | | Participants to telephone survey among non respondents | |
|---|---|---|---|---|---|---|
| **N** | 397 222 | | 22 496 | | 3 798 | |
| | N | % | N | % | N | % |
| **Age** (years) | | | | | | |
| Median age (IQR) | 42.0 (31.6 – 53.4) | | 45.4 (36.5 – 55.5) | | 42.0 (31.3 – 53.7) | |
| 18-39 | 178 832 | 45,0 | 7 696 | 34,2 | 1 820 | 45,8 |
| 40-49 | 93 038 | 23,4 | 6 358 | 28,3 | 884 | 22,2 |
| 50-59 | 77 829 | 19,6 | 5 094 | 22,6 | 775 | 19,5 |
| ≥60 | 47 523 | 12,0 | 3 348 | 14,9 | 499 | 12,5 |
| **Sex** (Female) | 309 300 | 77,9 | 18 151 | 80,7 | 3 113 | 78,3 |
| **Occupation**[a] | | | | | | |
| Physician | 18 450 | 4,6 | 1 178 | 5,2 | 177 | 4,4 |
| Nursing and respiratory care | 103 110 | 26,0 | 5 958 | 26,5 | 1 057 | 26,6 |
| Paratechnical personnel and auxiliary services | 110 885 | 27,9 | 3 728 | 16,6 | 1 062 | 26,7 |
| Administration staff | 63 558 | 16,0 | 3 694 | 16,4 | 655 | 16,7 |
| Health and social services technicians and professionals | 75 763 | 19,1 | 5 903 | 26,2 | 752 | 18,9 |
| Manager | 12 142 | 3,1 | 1 257 | 5,6 | 128 | 3,2 |
| Pharmacist | 5 574 | 1,4 | 237 | 1,1 | 69 | 1,7 |
| Others / Unknown | 7 740 | 1,9 | 541 | 2,4 | 68 | 1,7 |
| **Comorbidities**[b] | | | | | | |
| At least one comorbidity | 161 265 | 40,6 | 9 563 | 42,5 | 1 653 | 41,6 |
| At least two comorbidities | 60 213 | 15,2 | 3 672 | 16,3 | 591 | 14,9 |
| **COVID-19 vaccination at survey date** | | | | | | |
| 0 dose | 1 130 | 0,3 | 172 | 0,8 | 2 | 0,1 |
| 1 dose | 5 885 | 1,5 | 158 | 0,7 | 46 | 1,2 |
| 2 doses | 109 203 | 27,5 | 3 179 | 14,1 | 1 044 | 26,2 |
| 3 doses or more | 281 004 | 70,7 | 18 987 | 84,4 | 2 886 | 72,5 |
| **Sociosanitary region** | | | | | | |
| Montréal | 84 001 | 21,1 | 5 119 | 22,8 | 754 | 19,0 |
| Montérégie | 67 099 | 16,9 | 4 004 | 17,8 | 650 | 16,3 |
| Capitale-Nationale | 41 217 | 10,4 | 2 745 | 12,2 | 422 | 10,6 |
| Laurentides | 29 825 | 7,5 | 1 626 | 7,2 | 312 | 7,8 |
| Lanaudière | 27 992 | 7,0 | 1 311 | 5,8 | 294 | 7,4 |
| Mauricie et Centre-du-Québec | 26 455 | 6,7 | 1 347 | 6,0 | 284 | 7,1 |
| Estrie | 25 304 | 6,4 | 1 580 | 7,0 | 281 | 7,1 |
| Chaudières-Appalaches | 20 255 | 5,1 | 1 101 | 4,9 | 206 | 5,2 |
| Laval | 19 103 | 4,8 | 950 | 4,2 | 163 | 4,1 |
| Others (<4.5 %)[c] | 55 971 | 14,1 | 2 713 | 12,1 | 612 | 15,4 |

[a] Occupations based on administrative data, as self-reported occupation was not asked in the telephone survey

[b] At least two chronic conditions among the following: alcohol abuse, anaemia, cancer, cardiovascular disease, chronic respiratory disease, coagulopathy, diabetes, drug abuse, fluid and electrolyte disorders, history of depression, hypertension, hypothyroidism, kidney disease, liver disease, neurological disorder, obesity, psychosis, ulcer, paralysis, weight loss.

[c] Bas-Saint-Laurent, Saguenay-Lac-Saint-Jean, Outaouais, Abitibi-Témiscamingue, Côte-Nord, Nord-du-Québec, Gaspésie-Îles-de-la-Madeleine, Nunavik, Terres-Cries-de-la-Baie-James

Note: numbers in blue correspond to statistical differences ($p<0.05$) with the target population.

### eTable2. Long COVID cases and total COVID-19 cases by number of infections and episode of attribution of long COVID

| | Number of COVID-19 infections | | | | |
|---|---|---|---|---|---|
| | 1 | 2 | 3 | 4 | Total |
| **Cumulative risk:** | | | | | |
| COVID cases (n) | 12273 | 3755 | 576 | 57 | 16661 |
| Long COVID cases (n) | 1687 | 865 | 213 | 31 | 2796 |
| Cumulative long COVID risk (%, 95%CI) | 13.7% (13.1–14.4) | 23.0% (21.7–24.4) | 37.0% (33.0–40.9) | | 16.8% (16.1–17.2) |
| Episode of attribution of long COVID (n, %) | | | | | |
| 1st episode | 1687 (100.0) | 593 (68.8) | 147 (69.0) | 21 (67.7) | 2448 (87.6) |
| 2nd episode | 0 (0.0) | 179 (20.7) | 25 (11.7) | 1 (3.2) | 205 (7.3) |
| 3rd episode | 0 (0.0) | 0 (0.0) | 21 (9.9) | 1 (3.2) | 22 (0.8) |
| Unknown (or 4th episode) | 0 (0.0) | 93 (10.8) | 20 (9.4) | 8 (3.2) | 121 (4.3) |
| **Risk after 1st episode:** | | | | | |
| COVID cases (n) | 12273 | 3662 | 556 | 56 | 16547 |
| Long COVID cases (n) | 1687 | 593 | 147 | 21 | 2448 |
| Long COVID risk attributed to 1st episode (%, 95%CI) | | | | | 14.8% (14.3–15.3) |
| **Risk after 2nd episode:** | | | | | |
| COVID cases (n) | 0 | 3069 | 409 | 35 | 3513 |
| Long COVID cases (n) | 0 | 179 | 25 | 1 | 205 |
| Long COVID risk attributed to 2nd episode (%, 95%CI) | | | | | 5.8% (5.1–6.6) |
| **Risk after 3rd episode:** | | | | | |
| COVID cases (n) | 0 | 0 | 384 | 34 | 418 |
| Long COVID cases (n) | 0 | 0 | 21 | 1 | 22 |
| Long COVID risk attributed to 3rd episode (%, 95%CI) | | | | | 5.3% (3.1–7.4) |

Note: For each episode, contributing COVID cases (denominator) are total number of COVID cases minus long COVID cases with unknown attribution and minus long COVID cases occurring after a prior episode (e.g. COVID cases contributing to 2nd episode risk among HCWs with 2 infections: 3755 HCWs with 2 infections – 93 long COVID cases with unknown attribution – 593 long COVID cases attributed to 1st episode = 3069 COVID cases)

**eTable 3. Long COVID cases and total Omicron cases by number of infections and episode of attribution of long COVID, among HCWs who had only Omicron infections**

| | Number of Omicron infections | | | | |
|---|---|---|---|---|---|
| | 1 | 2 | 3 | 4 | Total |
| Cumulative risk: | | | | | |
| Omicron cases (n) | 10562 | 1969 | 163 | 6 | 12700 |
| Long COVID cases (n) | 1313 | 364 | 48 | 3 | 1728 |
| Cumulative long COVID risk (%, 95%CI) | 12.4% (11.8–13.1) | 18.5% (16.8–20.2) | 29.4% (22.5–36.4) | | 13.6% (13.0–14.2) |
| Episode of attribution of long COVID (n, %) | | | | | |
| 1st episode | 1313 (100.0) | 223 (61.3) | 34 (70.8) | 2 (66.7) | 1572 (91.0) |
| 2nd episode | 0 (0.0) | 92 (25.3) | 5 (10.4) | 0 (0.0) | 97 (5.6) |
| 3rd episode | 0 (0.0) | 0 (0.0) | 4 (8.3) | 0 (0.0) | 22 (0.8) |
| Unknown (or 4th episode) | 0 (0.0) | 49 (13.5) | 5 (10.4) | 1 (33.3) | 4 (0.2) |
| Risk after 1st episode: | | | | | |
| COVID cases (n) | 10562 | 1920 | 158 | 6 | 12646 |
| Long COVID cases (n) | 1313 | 315 | 43 | 2 | 1572 |
| Long COVID risk attributed to 1st episode (%, 95%CI) | | | | | 12.4% (11.8–13.1) |
| Risk after 2nd episode: | | | | | |
| COVID cases (n) | 0 | 1697 | 124 | 4 | 1825 |
| Long COVID cases (n) | 0 | 92 | 5 | 0 | 97 |
| Long COVID risk attributed to 2nd episode (%, 95%CI) | | | | | 5.3% (4.3–6.3) |
| Risk after 3rd episode: | | | | | |
| COVID cases (n) | 0 | 0 | 119 | 4 | 123 |
| Long COVID cases (n) | 0 | 0 | 4 | 0 | 4 |
| Long COVID risk attributed to 3rd episode (%, 95%CI) | | | | | 3.3% (0.1–6.4) |

Note: For each episode, contributing COVID cases (denominator) are total number of COVID cases minus long COVID cases with unknown attribution and minus long COVID cases occurring after a prior episode

### eTable 4. Disease severity, self-rated health and physical and cognitive impairment during daily activities, after exclusion of participants reporting all symptoms

| | Long COVID Cases | | | | COVID Controls | Non-COVID Controls |
|---|---|---|---|---|---|---|
| Characteristic | Overall | Mild | Moderate | Severe | | |
| **N** | **1888** | **512** | **844** | **532** | **11697** | **4755** |
| **Reported symptoms, n (%)** | | | | | | |
| Median (IQR) | 4 (2 – 8) | 2 (1 – 4) | 5 (3 – 8) | 9 (5 – 13) | 0 (0 – 2) | 0 (0 – 3) |
| None | 0 (0.0) | NA | NA | NA | 7588 (64.9) | 2504 (52.7) |
| 1 – 2 | 573 (30.3) | 303 (59.2) | 207 (24.5) | 63 (11.8) | 1827 (15.6) | 816 (17.2) |
| 3 – 4 | 374 (19.8) | 121 (23.6) | 199 (23.6) | 54 (10.2) | 1027 (8.8) | 591 (12.4) |
| 5 – 9 | 563 (29.8) | 85 (16.6) | 312 (37.0) | 166 (31.2) | 999 (8.5) | 646 (13.6) |
| 10 – 14 | 271 (14.4) | 2 (0.4) | 104 (12.3) | 165 (31.0) | 136 (1.2) | 153 (3.2) |
| 15 – 19 | 107 (5.7) | 1 (0.2) | 22 (2.6) | 84 (15.8) | 120 (1.0) | 45 (1.0) |
| **Reported moderate or severe symptoms, n (%)** | | | | | | |
| Median (IQR) | 1 (0 – 4) | NA | 2 (1 – 3) | 6 (3 – 9) | 0 (0 – 0) | 0 (0 – 1) |
| None | 512 (27.1) | NA | NA | NA | 9460 (80.9) | 3370 (70.9) |
| 1 – 2 | 655 (34.7) | NA | 534 (63.3) | 121 (22.7) | 1305 (11.2) | 697 (14.7) |
| 3 – 4 | 264 (14.0) | NA | 177 (21.0) | 87 (16.4) | 481 (4.1) | 324 (6.8) |
| 5 – 9 | 339 (18.0) | NA | 124 (14.7) | 215 (40.4) | 392 (3.4) | 303 (6.4) |
| 10 – 14 | 101 (5.3) | NA | 9 (1.1) | 92 (17.3) | 50 (0.4) | 54 (1.1) |
| 15 – 19 | 17 (0.9) | NA | 0 (0.0) | 17 (3.2) | 9 (0.1) | 7 (0.2) |
| **Cluster (symptoms)[a], n (%)** | | | | | | |
| C1-Systemic (fatigue, PEM, headache, muscular/joint pain) | 1470 (77.9) | 288 (56.3) | 688 (81.5) | 494 (92.9) | 3349 (28.6) | 1888 (39.7) |
| C2-Neurocognitive (difficulty with concentration, forgetfulness, brain fog) | 1044 (55.3) | 178 (34.8) | 471 (55.8) | 395 (74.2) | 1577 (13.5) | 969 (20.4) |
| C3-Pulmonary (shortness of breath) | 915 (48.5) | 169 (33.0) | 417 (49.4) | 329 (61.8) | 550 (4.7) | 254 (5.3) |
| C4-Mood (anxiety. depression. sleep problems) | 872 (46.2) | 99 (19.3) | 395 (46.8) | 378 (71.1) | 2071 (7.7) | 1446 (30.4) |
| C5 Upper respiratory (cough, runny nose, sore throat, tightness of chest, fever) | 785 (41.6) | 160 (31.3) | 326 (38.6) | 299 (56.2) | 1352 (11.6) | 733 (15.4) |
| C6-Smell/taste impairment | 512 (27.1) | 112 (21.9) | 219 (25.9) | 181 (34.0) | 217 (1.9) | 92 (1.9) |
| C7 Digestive (abdominal pain, diarrhea) | 284 (15.0) | 14 (2.7) | 93 (11.0) | 177 (33.3) | 454 (3.9) | 272 (5.7) |
| **Self-rated health, n (%)** | | | | | | |
| Excellent / Very good | 103 (5.5) | 62 (12.1) | 38 (4.5) | 3 (0.6) | 4021 (34.4) | 1368 (28.8) |
| Good / Fair | 1286 (68.1) | 414 (80.9) | 637 (75.5) | 235 (44.0) | 7087 (60.6) | 2959 (62.2) |
| Poor | 499 (26.4) | 36 (7.0) | 169 (20.0) | 294 (55.3) | 589 (5.0) | 428 (9.0) |
| **Effort performance (mMRC score)[b], n (%)** | | | | | | |

| | | | | | | |
|---|---|---|---|---|---|---|
| Moderate limitation | 1496 (79.2) | 365 (71.3) | 676 (80.1) | 455 (85.5) | 4964 (42.4) | 2127 (44.7) |
| Severe limitation | 28 (1.5) | 1 (0.2) | 7 (0.8) | 20 (5.3) | 14 (0.1) | 17 (0.4) |
| **Cognitive impairment[c]** | | | | | | |
| Difficulty to concentrate | 627 (33.2) | 80 (15.6) | 272 (32.2) | 275 (51.7) | 1228 (10.5) | 574 (12.1) |
| Forgetfulness | 571 (30.2) | 72 (14.1) | 249 (29.5) | 250 (47.0) | 1176 (10.1) | 491 (10.3) |
| Difficulty to organize oneself | 428 (22.7) | 51 (10.0) | 174 (20.6) | 203 (38.2) | 585 (7.3) | 422 (8.9) |
| Loss of necessary items | 322 (17.1) | 35 (6.8) | 139 (16.5) | 148 (27.8) | 617 (5.3) | 281 (5.9) |

[a] Presence of at least one symptom of each cluster

[b]mMRC score: moderate limitation = shortness of breath on level ground, score 3-4/5 and severe limitation = shortness of breath at minimal effort, score 5/5

[c] Cognitive impairment defined as reporting the listed difficulties “often or very often” during daily activities

Abbreviations: IQR, interquartile range; mMRC scale, Modified Medical Research Council Dyspnea Scale

### eTable 5. Symptom duration of long COVID cases after first (and single) COVID-19 episode

| | Prevalent long COVID cases | | | | Recovered long COVID cases |
|---|---|---|---|---|---|
| | Overall[a] | Mild | Moderate | Severe | |
| N | 1329 | 333 | 567 | 387 | 151 |
| Mean duration in months (STD) | 15.1 (9.2) | 13.5 (8.4) | 15.1 (9.3) | 16.4 (9.7) | 7.3 (6.1) |
| Median duration in months (IQR) | 12.6 (8.9-17.1) | 11.3 (8.0-15.8) | 12.5 (8.5-17.3) | 13.6 (10.0-18.5) | 4.7 (3.3-9.6) |
| Duration | | | | | |
| 3-5 months | 152 (11.4) | 47 (14.1) | 65 (11.5) | 35 (9.0) | 93 (61.6) |
| 6 -11 months | 484 (36.4) | 138 (41.4) | 208 (36.7) | 122 (31.5) | 29 (19.2) |
| 12-23 months | 439 (33.0) | 101 (30.3) | 183 (32.3) | 139 (35.9) | 23 (15.2) |
| 24-35 months | 171 (12.9) | 32 (9.6) | 76 (13.4) | 59 (15.2) | 5 (3.3) |
| ≥36 months | 83 (6.2) | 15 (4.5) | 35 (6.2) | 32 (8.3) | 1 (0.7) |

[a] Including 42 cases with no information on symptom severity

Abbreviations: IQR, interquartile range; STD, standard deviation
